# Angiotensin II Receptor Blockers and Angiotensin-Converting Enzyme Inhibitors Usage is Associated with Improved Inflammatory Status and Clinical Outcomes in COVID-19 Patients With Hypertension

**DOI:** 10.1101/2020.03.31.20038935

**Authors:** Guang Yang, Zihu Tan, Ling Zhou, Min Yang, Lang Peng, Jinjin Liu, Jingling Cai, Ru Yang, Junyan Han, Yafei Huang, Shaobin He

## Abstract

With the capability of inducing elevated expression of ACE2, the cellular receptor for SARS-CoV-2, angiotensin II receptor blockers or angiotensin-converting enzyme inhibitors (ARBs/ACEIs) treatment may have a controversial role in both facilitating virus infection and reducing pathogenic inflammation. We aimed to evaluate the correlation of ARBs/ACEIs usage with the pathogenesis of COVID-19 in a retrospective, single-center study. 126 COVID-19 patients with preexisting hypertension at Hubei Provincial Hospital of Traditional Chinese Medicine (HPHTCM) in Wuhan from January 5 to February 22, 2020 were retrospectively allocated to ARBs/ACEIs group (n=43) and non-ARBs/ACEIs group (n=83) according to their antihypertensive medication. 125 age- and sex-matched COVID-19 patients without hypertension were randomly selected as non-hypertension controls. In addition, the medication history of 1942 hypertension patients that were admitted to HPHTCM from November 1 to December 31, 2019 before COVID-19 outbreak were also reviewed for external comparison. Epidemiological, demographic, clinical and laboratory data were collected, analyzed and compared between these groups. The frequency of ARBs/ACEIs usage in hypertension patients with or without COVID-19 were comparable. Among COVID-19 patients with hypertension, those received either ARBs/ACEIs or non-ARBs/ACEIs had comparable blood pressure. However, ARBs/ACEIs group had significantly lower concentrations of CRP (p=0.049) and procalcitonin (PCT, p=0.008). Furthermore, much lower proportion of critical patients (9.3% *vs* 22.9%; p=0.061), and a lower death rate (4.7% *vs* 13.3%; p=0.216) were observed in ARBs/ACEIs group than non-ARBs/ACEIs group, although these differences failed to reach statistical significance. Our findings thus support the use of ARBs/ACEIs in COVID-19 patients with preexisting hypertension.

## Introduction

In December, 2019, a novel coronavirus disease (COVID-19) caused by severe acute respiratory syndrome coronavirus 2 (SARS-CoV-2) emerged in Wuhan, Hubei province of China. The disease has rapidly spread from Wuhan to other areas. As of March 25, 2020, 414,179 cases have been reported in 180 countries and areas from 6 continents, with the current crude case fatality rate (CFR) of 4.5%.^1^ Thus, with accumulating cases and high CFR, COVID-19 has posed a great challenge to public health.

According to a recent report, 91.1% of patients infected with SARS-CoV-2 were diagnosed pneumonia during hospitalization, including 15.7% of them with severe disease.^2^ However, the underlying pathophysiological mechanisms by which this virus causes disease remain unclear. SARS-CoV-2 and SARS-CoV are both coronaviruses, and the two viruses share 79% identity in nucleotide sequence.^3^ Early studies has established that SARS-CoV uses angiotensin-converting enzyme 2 (ACE2) as cellular receptor to facilitate its infection.^4,5^ In light of this, a recent investigation predicted that the receptor binding domain (RBD) of the spike glycoprotein from SARS-CoV-2 and SARS-CoV share almost identical conformation, and the binding affinity between RBD and ACE2 was estimated to be much stronger for SARS-CoV-2, as assessed by computer modeling.^6^ This notion was later confirmed by biophysical analysis.^7^ Indeed, SARS-CoV-2 can use ACE2 but not other tested proteins as a cellular entry receptor in virus infectivity studies using HeLa cell expressing human ACE2.^8^ ACE2 is widely expressed in many human tissues such as lung, intestine, heart, kidney and endothelium, suggesting that these tissue may serve as entry sites for infection and replication for both SARS-CoV and SARS-CoV-2.^9^ Upon binding to ACE2, SARS-CoV subsequently induced the downregulation of ACE2 in host cells, resulting in increased expression of Ang II which in turn caused severe acute lung injury.^10,11^ Taken together, early investigations in SARS-CoV suggest that ACE2 may have both a pathogenic role in facilitating virus infection, and a protective effect in limiting lung injury during SARS-CoV-2 infection.^12^ Despite its implication in virus infection, ACE2 is initially recognized as a critical enzymes in the renin-angiotensin system (RAS) that regulates blood pressure, fluid and ecectrolyte balance, and vascular resistance.^13^ In fact, drugs that can upregulate the expression of ACE2 such as ARBs and ACEIs, have been extensively used in patients with hypertension and other cardiovascular diseases to maintain the stability of blood pressure and reduce the risk of adverse events in cardio-cerebrovascular system and kidney.^14-16^

Hypertension represented a major comorbidity in patients infected with SARS-CoV-2. According to recent reports in China and Singapore, 12.8% to 31.2% of COVID-19 patients had preexisting hypertension.^2,17-22^ These patients appeared to develop severe disease more frequently and were more susceptible to death.^2,19^ Since a number of these patients used ARBs/ACEIs as antihypertensive drugs, which can increase the expression of ACE2, and because of the seemly paradoxical role of ACE2 during SARS-CoV-2 infection, we designed this retrospective study to determine whether the usage of ARBs/ACEIs are associated with SARS-CoV-2 infection and the clinical outcomes in COVID-19 patients with preexisting hypertension.

## Methods

### Study Design and Participants

This retrospective study complied with the Declaration of Helsinki and was approved by the Hubei Provincial Hospital of Traditional Chinese Medicine (HPHTCM)’s ethical review board (Clinical Ethical Approval No. HBZY2020-C15-01). HPHTCM is responsible for the treatments of COVID-19 assigned by the Wuhan government. Patients with confirmed COVID-19 according to the guideline of SARS-CoV-2 (The Fifth Trial Version of the Chinese National Health Commission) admitted into HPHTCM from January 5 to February 22, 2020, were included for initial screen.^23^ COVID-19 patients with preexisting hypertension were retrospectively allocated into two subgroups, ARBs/ACEIs and non-ARBs/ACEIs group, according to their usage of antihypertensive drugs. Age- and sex-matched cases were randomly selected from the remaining COVID-19 patients without hypertension as non-hypertension controls. The clinical outcomes (ie, discharges, mortality, length of stay) were monitored up to March 3, 2020, the final date of follow-up. In addition, the medication history of 1942 hypertension patients that were admitted to HPHTCM from November 1 to December 31, 2019 before COVID-19 outbreak were also reviewed for external comparison. Written informed consent was waived by the Ethics Commission of the hospital for emerging infectious diseases.

### Data Collection

The medical records of patients were analyzed by the research team of HPHTCM. Disease onset was defined as the date when the symptom was noticed. Data on the use of ACEIs and ARBs prior to admission and during hospital stay were collected. Other information including demographic data, medical history, exposure history, comorbidities, symptoms, signs, laboratory findings, and treatment measures (ie, antiviral therapy, corticosteroid therapy) were extracted from electronic medical records and were recorded with standardized data collection forms. The data were reviewed by a trained team of physicians.

Assessment of disease status followed the guideline of SARS-CoV-2 (The Fifth Trial Version of the Chinese National Health Commission): mild type, with slight clinical symptoms but no imaging presentations of pneumonia; common type, with fever, respiratory tract and other symptoms, imaging findings of pneumonia; severe type, with any of the following conditions: respiratory distress, respiratory frequency ≥30 times/minutes, finger oxygen saturation at rest ≤93%, or oxygenation index [PaO_2_/ FiO_2_]≤300 mmHg (1 mmHg=0.133 kPa); critical type, with any of the following conditions: respiratory failure requires mechanical ventilation, shock, combined with other organ failure requires intensive care unit care and treatment.^21^ Laboratory parameters, including complete blood count, C-reactive protein (CRP), arterial blood gas analysis, myocardial injury markers, coagulation profile, serum biochemical tests (including renal and liver function, lactate dehydrogenase), procalcitonin (PCT), b-type natriuretic peptide (BNP), were measured according to the manufacturer’s instructions.

### Statistical Analysis

Data analysis was performed using SPSS (Statistical Package for the Social Sciences, version 23). Categorical variables were reported as absolute (relative frequencies) and compared by χ tests or Fisher’s exact tests. Continuous variables were expressed as mean (SD) if they are normally distributed or median (interquartile range, IQR) if they are not and compared by independent group t tests or Mann-Whitney U tests, respectively. p<0.05 was considered as statistically significant.

## Results

### Presenting Characteristics

After the initial screen, 462 COVID-2019 patients were allocated to two groups, the hypertension group that includes 126 patients with preexisting hypertension, and the non-hypertension group comprising 125 age- and sex-matched patients that were randomly selected from the remaining patients without hypertension (figure 1). The median age of the hypertension group was 66 (IQR, 61-73) years, and 62 (49.2%) were men, which is similar compared to 66 (IQR, 60-75) years and 61 (48.8%) in the non-hypertension group. Compared with non-hypertension controls, patients with preexisting hypertension had higher levels of systolic blood pressure (125 [IQR, 120-140] *vs* 120 [IQR, 120-134]; p=0.031) and diastolic blood pressure(75 [IQR, 70-85] *vs* 70 [IQR, 70-80]; p=0.004), as well as higher proportion of diabetes (30.2% [38 of 126] *vs* 13.6% [17 of 125]; p=0.002) and cardiopathy (18.3% [23 of 126] *vs* 9.6% [12 of 125]; p=0.048).

**Figure 1.**
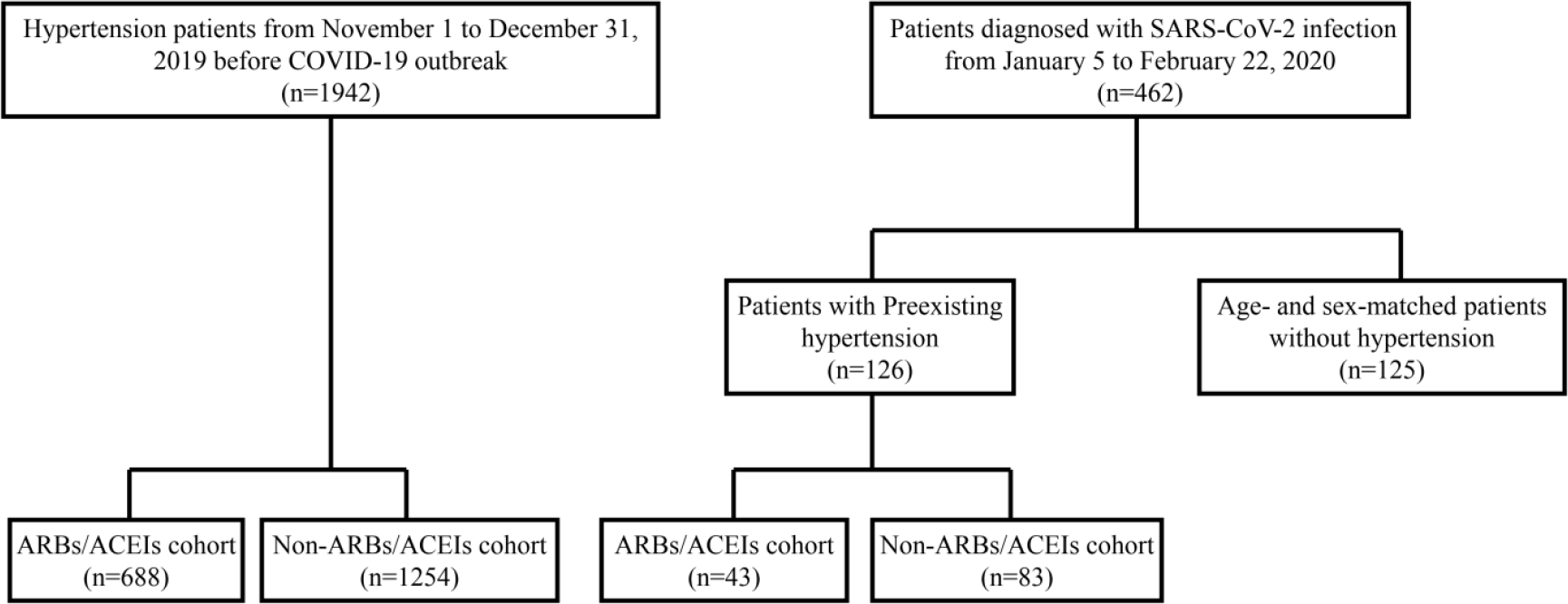
Flowchart of the selection for study subjects. Abbreviations: ACEIs, angiotensin-converting enzyme inhibitors; ARBs, angiotensin II receptor blockers.

COVID-19 patients with hypertension was further allocated into two subgroups based on the usage of ARBs/ACEIs as antihypertensive drugs: 43 in the ARBs/ACEIs subgroup and 83 in the non-ARBs/ACEIs subgroup (figure 1). The median ages of patients in the two subgroups were 67 (IQR, 62-75) and 65 (IQR, 57-72) years, respectively. There were no major differences in characteristics between the two subgroups except for the higher usage of antibiotics in the non-ARBs/ACEIs subgroup. Baseline characteristics are shown in table 1. The age distribution and complication ratio of hypertension group and non-hypertension group is shown in figure 2.

**Table 1.** **Demographics, baseline characteristics, and complications of 126 hypertension patients and 125 non-hypertension patients with COVID-19 at Hubei Provincial Hospital of Traditional Chinese Medicine (Jan 5 to Feb 22, 2020)**.

|  | Non-Hypertension |  | Hypertension |  | *P* Value | *P* Value |
| --- | --- | --- | --- | --- | --- | --- |
|  | Total (n=125) | Total (n=126) | Non-ARBs/ACEIs<br>(n=83) | ARBs/ACEIs<br>(n=43) | Non-Hypertension<br>vs Hypertension | Non-ARBs/ACEIs<br>vs ARBs/ACEIs |
| **Age, median (IQR)** | 66 (60-72) | 66 (61-73) | 67 (62-75) | 65 (57-72) | 0.460 | 0.122 |
| **Sex , No. (%)** |  |  |  |  | 0.949 | 0.952 |
| Female | 64 (51.2) | 64 (50.8) | 42 (50.6) | 22 (51.2) |  |  |
| Male | 61 (48.8) | 62 (49.2) | 41 (49.4) | 21 (48.8) |  |  |
| **Anthropometrics, median (IQR)** |  |  |  |  |  |  |
| Weight (Kg) | 60.0 (56.5-65.0) | 60.0 (57.5-70.0) | 60.0 (55.0-65.0) | 60.0 (60.0-70.0) | 0.516 | 0.179 |
| Height (cm) | 165.0<br>(160.0-170.0) | 165.0<br>(160.0-171.0) | 162.0 (159.0-170.0) | 166.0<br>(160.0-173.0) | 0.735 | 0.345 |
| BMI (Kg/m<sup>2</sup>) | 23.25<br>(20.28-24.20) | 22.86<br>(20.83-24.77) | 22.77 (20.81-24.61) | 23.42<br>(21.09-25.39) | 0.525 | 0.305 |
| **Blood pressure (mmHg) , median (IQR)** |  |  |  |  |  |  |
| Systolic blood pressure | 120 (120-134) | 125 (120-140) | 124 (120-143) | 129 (120-140) | **0.031** | 0.966 |
| Diastolic blood pressure | 70 (70-80) | 75 (70-85) | 75 (70-85) | 77 (70-85) | **0.004** | 0.688 |
| **Symptom, No. (%)** |  |  |  |  |  |  |
| Fever | 84 (67.2) | 90 (71.4) | 60 (72.3) | 30 (69.8) | 0.468 | 0.766 |
| Cough | 84 (67.2) | 87 (69.0) | 57 (68.7) | 30 (69.8) | 0.753 | 0.900 |
| Headache | 15 (12.1) | 20 (15.9) | 12 (14.5) | 8 (18.6) | 0.376 | 0.546 |
| Durchfall | 13 (10.5) | 19 (15.1) | 13 (15.7) | 6 (14.0) | 0.266 | 0.779 |
| **Complications, No. (%)** |  |  |  |  |  |  |
| Diabetes | 17 (13.6) | 38 (30.2) | 25 (30.1) | 13 (30.2) | **0.002** | 0.990 |
| Respiratory disease | 6 (4.4) | 6 (4.7) | 3 (3.6) | 3 (7.0) | 0.903 | 0.690 |
| Kidney disease | 1 (0.8) | 3 (2.4) | 3 (3.6) | 0 (0.0) | 0.317 | 0.207 |
| Hepatic disease | 5 (4.1) | 8 (6.3) | 5 (6.1) | 3 (7.0) | 0.401 | 0.849 |
| Cardiopathy | 12 (9.6) | 23 (18.3) | 16 (19.3) | 7 (16.3) | **0.048** | 0.680 |
| Neurological disease | 5 (4.0) | 10 (7.9) | 6 (7.2) | 4 (9.3) | 0.188 | 0.683 |
| Immune diseases | 0 (0.0) | 0 (0.0) | 0 (0.0) | 0 (0.0) | - | - |
| **Treatments, No. (%)** |  |  |  |  |  |  |
| Glucocorticoid | 35 (28.0) | 39 (31.0) | 23 (27.7) | 16 (37.2) | 0.608 | 0.274 |
| Antiviral | 100 (80.0) | 96 (76.2) | 66 (79.5) | 30 (69.8) | 0.466 | 0.223 |
| Antibiotic | 74 (59.2) | 98 (77.8) | 69 (83.1) | 29 (67.4) | **0.002** | **0.045** |
| Immunoglobulin | 28 (24.1) | 40 (31.7) | 25 (30.1) | 15 (34.9) | 0.151 | 0.586 |

Data are median (IQR) or n (%). *P* values were obtained from $\chi^2$ test, Fisher's exact tests, T tests or Mann-Whitney U tests, when appropriate. *P* <0.05 was considered statistically significant (**in bold**).

**Figure 2.**
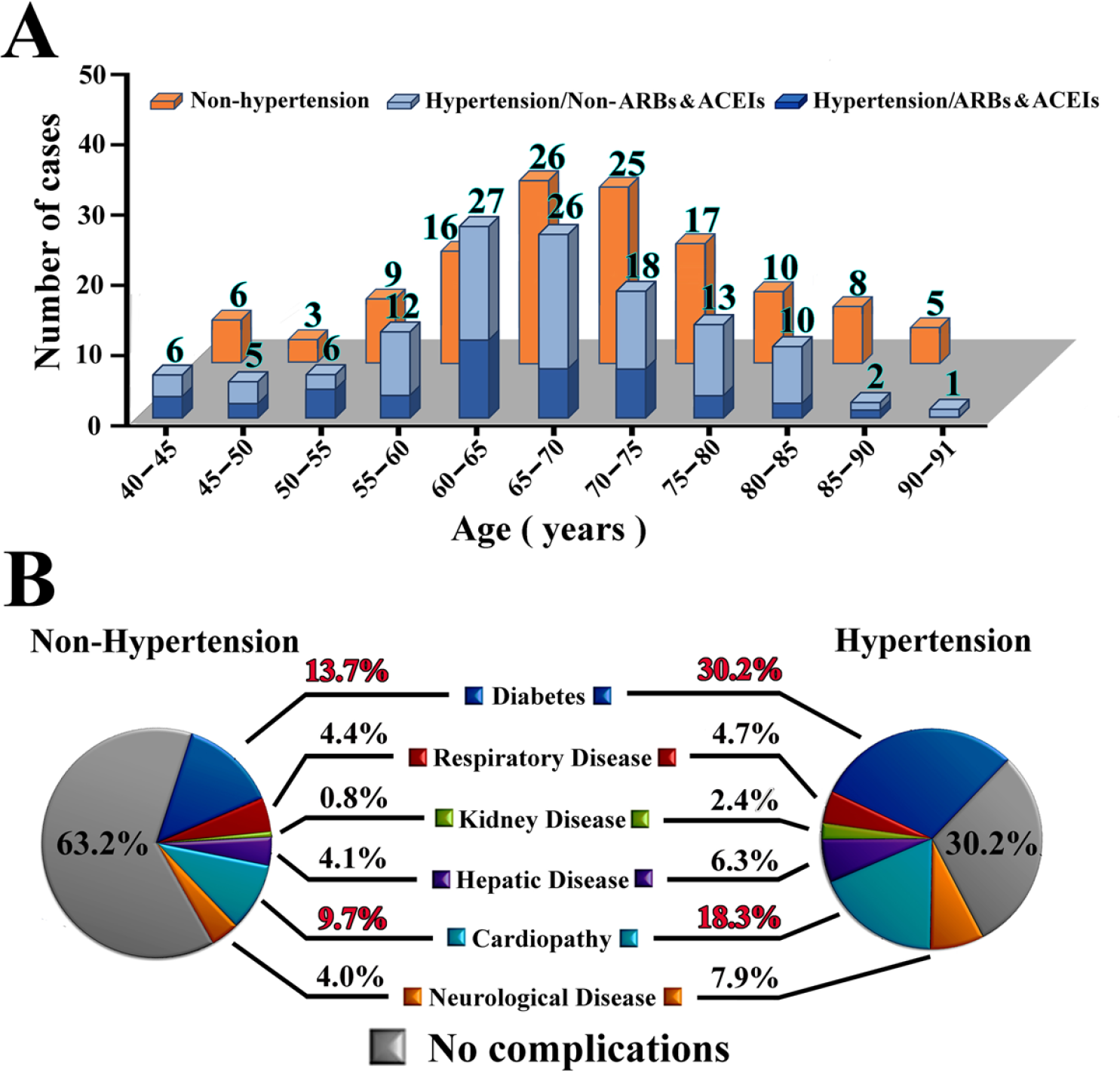
Age distribution and the frequencies of complications in hypertension group and non–hypertension group.

Of note, the frequency of ARBs/ACEIs usage in COVID-2019 patients with hypertension was 34.1% (43 of 126), which is not statistically different from that observed in hypertension patients admitted to HPHTCM before COVID-19 emerged (35.4% [688 of 1952]; p=0.767), and the age/sex distributions are comparable between these two groups (figure 3).

**Figure 3.**
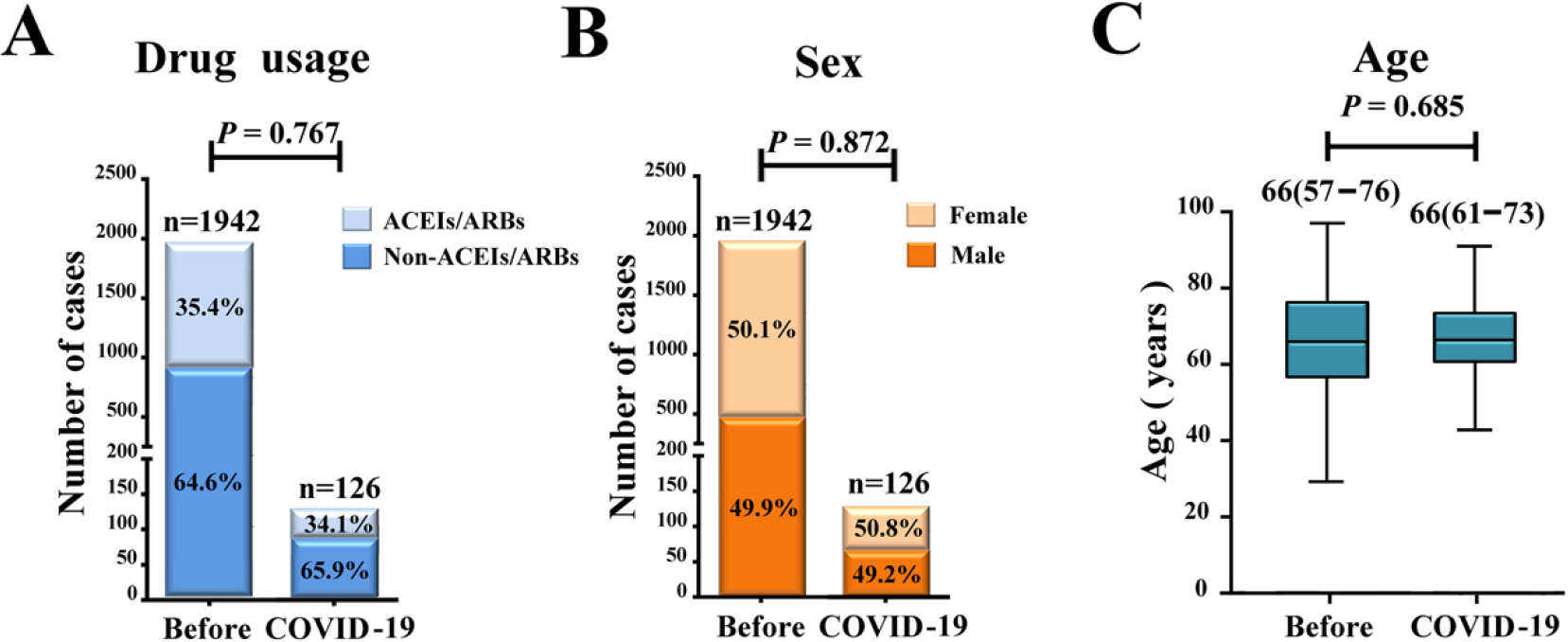
Usage of antihyperhensive drugs (A), sex ratio (B), and age distribution (C) of hypertension patients with or without COVID-19. Without COVID-19 (before): patients admitted to Hubei Provincial Hospital of Traditional Chinese Medicine before COVID-19 outbreak (from Nov 1 to Dec 31, 2019).

### Disease Status, Course and Outcome

Among the 126 patients with hypertension, 23 (18.3%) were critical type, 27 (21.4%) were severe, and 76 (60.3%) were mild and common. During the observation period, 13 (10.3%) patients in the hypertension group died, 71 (56.3%) discharged, and 42 (33.3%) remained in hospital (table 2). Compared with the hypertension group, the non-hypertension group seemingly had fewer cases of critical illness (14 [11.2%]) and fewer deaths (8 [6.4%]), but this difference is not statistically significant. Those with preexisting hypertension had longer disease courses for not recovered patients (36.7±12.4 *vs* 31.6±7.7; p=0.024).

**Table 2.** **Classification of severity, outcome and disease course of hypertension and non-hypertension patients with COVID-19**.

|  | Non-Hypertension |  | Hypertension |  | *P* Value | *P* Value |
| --- | --- | --- | --- | --- | --- | --- |
|  | Total(n=125) | Total (n=126) | Non-ARBs/ACEIs (n=83) | ARBs/ACEIs (n=43) | Non-Hypertension vs Hypertension | Non-ARBs/ACEIs vs ARBs/ACEIs |
| **Classification of severity No. (%)** |  |  |  |  | 0.156 | 0.162 |
| Mild/ Common | 89 (71.2) | 76 (60.3) | 48 (57.8) | 28 (65.1) |  |  |
| Severe | 22 (17.6) | 27 (21.4) | 16 (19.3) | 11 (25.6) |  |  |
| Critical | 14 (11.2) | 23 (18.3) | 19 (22.9) | 4 (9.3) | 0.115 | 0.061 |
| **Outcome No. (%)** |  |  |  |  | 0.523 | 0.283 |
| Discharged | 72 (57.6) | 71 (56.3) | 44 (53.0) | 27 (62.8) |  |  |
| Remained in hospital | 45 (36.0) | 42 (33.3) | 28 (33.7) | 14 (32.6) |  |  |
| Died | 8 (6.4) | 13 (10.3) | 11 (13.3) | 2 (4.7) | 0.383 | 0.216 |
| **Disease course (Days), Mean, SDs** |  |  |  |  | 0.501 | 0.880 |
| Discharged | 28.6±8.0 | 29.6±12.6 | 28.4±10.0 | 25.6±9.5 | 0.811 | 0.227 |
| Remained in hospital | 31.6±7.7 | 36.7±12.4 | 37.5±12.3 | 35.2±12.8 | **0.024** | 0.597 |
| Died | 20.3±5.7 | 15.4±9.9 | 14.7±10.7 | 19.0±1.4 | 0.223 | 0.598 |

Data are n (%) or mean ± SDs. *P* values were obtained from $\chi^2$ tests, Fisher's exact tests, or Mann-Whitney U tests, when appropriate. *P* < 0.05 was considered statistically significant (**in bold**).

Furthermore, within the hypertension group, patients on ARBs/ACEIs had a much lower proportion of critical patients (9.3% [four of 43] *vs* 22.9% [19 of 83]; p=0.061), and a lower death rate (4.7% [two of 43] *vs* 13.3% [11 of 83]; p=0.283) than those on non-ARBs/ACEIs medications, although these differences failed to reach statistical significance (table 2).

### 3. Laboratory Testing

Compared with non-hypertensive controls, COVID-19 patients with preexisting hypertension had lower arterial partial pressure of oxygen (10.1 [7.6-11.3] *vs* 11·3 [8·9-13·8]; p=0·001), oxygen index (95 [90-97] *vs* 97 [94-98]; p<0.001), higher blood urea (5.0 [3.8-8.5] *vs* 4.5 [3.5-5.5], p=0.020), ALT(28 [17-55] v*s* 22 [14-43]; p=0.022) and cardiac troponin (0.006 [0-0.031] *vs* 0 [0-0.014]; p=0.015), and higher concentrations of hypersensitive-CRP (25.4 [4.6-100.8] *vs* 12.6 [2.6-53.3]; p=0.024), procalcitonin (0.092 [0.049-0.223] *vs* 0.062 [0.035-0.134]; p=0.017) and IL-6 (13.8 [4.8-51.3] *vs* 8.2 [1.8-22.8]; p=0.017). In COVID-19 patients with preexisting hypertension, ARBs/ACEIs treatment significantly reduced the concentrations of CRP (11.5 [4.0-58.2] *vs* 33.9 [5.1-119.2]; p=0.049) and procalcitonin (0.061 [0.044-0.131] *vs* 0.121 [0.052-0.295]; p=0.008), when compared with non-ARBs/ACEIs treatment (table 3).

**Table 3.** Laboratory findings of patients infected with SARS-CoV-2 at admission to HPHTCM.

|  | Normal range | Non-Hypertension |  | Hypertension |  | P Value | P Value |
| --- | --- | --- | --- | --- | --- | --- | --- |
|  |  | Total(n=125) | Total (n=126) | Non-ARBs/ACEIs<br>(n=83) | ARBs/ACEIs<br>(n=43) | Non-Hyper<br>tension vs<br>Hypertensi<br>on | Non-ARB<br>s/ACEIs<br>vs<br>ARBs/AC<br>EIs |
| Blood routine, median (IQR) |  |  |  |  |  |  |  |
| Neutrophil count | 1.8-6.3 ×10<sup>9</sup>/L | 3.2 (2.4-5.2) | 4.1 (2.9-6.7) | 4.2 (3.0-7.0) | 4.0 (2.9-6.3) | 0.001 | 0.934 |
| Lymphocyte count | 1.1-3.2 ×10<sup>9</sup>/L | 1.13 (0.76-1.46) | 0.99 (0.63-1.47) | 1.03 (0.74-1.47) | 0.84 (0.51-1.39) | 0.204 | 0.296 |
| Blood oxygen index, median (IQR) |  |  |  |  |  |  |  |
| Arterial partial pressure of oxygen | 10.6-13.3 KPa | 11.3 (8.9-13.8) | 10.1 (7.6-11.3) | 10.2 (7.2-11.1) | 10.1 (8.2-12.2) | 0.001 | 0.248 |
| Oxygen saturation | 95-98% | 97 (94-98) | 95 (90-97) | 95 (89-97) | 95 (91-97) | 0.000 | 0.812 |
| Coagulation marker, median (IQR) |  |  |  |  |  |  |  |
| D-dimer, mg/L | 0-0.50 mg/L | 0.41 (0.28-0.94) | 0.42 (0.29-1.34) | 0.47 (0.29-1.82) | 0.40 (0.30-0.61) | 0.895 | 0.284 |
| Prothrombin time | 10-14 s | 12.2 (11.5-12.7) | 12.2 (11.6-13.0) | 12.3 (11.6-13.1) | 12.1 (11.4-12.9) | 0.476 | 0.257 |
| Blood biochemistry, median(IQR) |  |  |  |  |  |  |  |
| Urea | 3.6-9.5 μmol/L | 4.5 (3.5-5.5) | 5.0 (3.8-8.5) | 5.1 (3.6-6.4) | 4.8 (3.9-6.0) | 0.020 | 0.751 |
| Creatinine | 57-110 μmol/L | 65 (55-79) | 69 (58-88) | 69 (59-87) | 69 (58-90) | 0.080 | 0.987 |
| Glomerular filtration rate | >90 mL/Min.1.73m<sup>2</sup> | 105.6 (88.2-128.4) | 101.8 (79.2-120.3) | 102.1 (78.2-119.2) | 101.4 (80.0-121.0) | 0.096 | 0.927 |
| Direct bilirubin | 0-4 μmol/L | 3.0 (2.2-4.2) | 3.2 (2.3-4.6) | 3.1 (2.4-4.6) | 3.2 (2.1-5.0) | 0.226 | 0.821 |
| Indirect bilirubin | 0-19 μmol/L | 5.6 (4.0-8.3) | 5.8 (4.4-8.5) | 5.6 (4.4-8.2) | 6.1 (4.8-9.1) | 0.443 | 0.454 |
| Alanine transaminase (ALT) | 9-50 U/L | 22 (14-43) | 28 (17-55) | 26 (17-53) | 34 (18-58) | **0.022** | 0.620 |
| Aspartate transferase (AST) | 15-40 U/L | 21 (17-35) | 24 (17-38) | 24 (18-39) | 23 (17-31) | 0.344 | 0.551 |
| Cardiac troponin | 0-0.06 ng/mL | 0 (0-0.014) | 0.006 (0-0.031) | 0.0075 (0-0.0493) | 0 (0-0.014) | **0.015** | 0.074 |
| Pro-brain natriuretic peptide (Pro-BNP) | 0-125.2 pg/mL | 174.7 (75.3-597.4) | 219.1 (92.0-1151.3) | 236.9 (72.0-3256.0) | 195.1 (106.2-651.4) | 0.153 | 0.354 |
| Creatine kinase MB | 0-5 ng/mL | 0.88 (0.56-1.63) | 0.94 (0.65-1.73) | 0.93 (0.65-1.62) | 1.09 (0.60-2.35) | 0.369 | 0.571 |
| Lactate dehydrogenase | 0-25 U/L | 200 (152-251) | 222 (182-302) | 228 (188-323) | 202 (179-258) | 0.018 | 0.125 |
| **Infection-related biomarkers, median (IQR)** |  |  |  |  |  |  |  |
| Hypersensitive C-reactive protein | 0-3mg/L | 12.6 (2.6-53.3) | 25.4 (4.6-100.8) | 33.9 (5.1-119.2) | 11.5 (4.0-58.2) | **0.024** | **0.049** |
| Procalcitonin | 0-0.052 ng/mL | 0.062 (0.035-0.134) | 0.092 (0.049-0.223) | 0.121 (0.052-0.295) | 0.061 (0.044-0.131) | **0.017** | **0.008** |
| Interleukin-6 | <7 pg/mL | 8.2 (1.8-22.8) | 13.8 (4.8-51.3) | 14.3 (3.7-121.1) | 10.1 (5.0-50.4) | **0.017** | 0.520 |

Data are median (IQR). *P* values were from T tests or Mann-Whitney U tests. *P* <0.05 was considered statistically significant (**in bold**).

## Discussion

This retrospective investigation, to our knowledge, is the first case-control study aimed to examine the correlation of ARBs/ACEIs between the pathogenesis of SARS-CoV-2 infection in patients with preexisting hypertension. The major finding of this report is that ARBs/ACEIs treatment was associated with reduced inflammatory response and mitigated disease progression in COVID-19 patients with preexisting hypertension compared to other antihypertensive treatment.

Hypertension is the leading cause of mortality globally. 31.2% of adults were estimated to have hypertension worldwide in 2010.^24^ Since the emerge of SARS-CoV-2 infection, hypertension has been frequently observed as a major comorbidity in COVID-19 patients.^2,17,20-22^ We reported here that 27.2% (126 of 462) of the COVID-19 patients admitted to HPHTCM have preexisting hypertension. This incidence is similar to those reported by Young et al (28%), Zhang et al (30%) and Wang et al (31.2%),^20-22^ but higher than that reported in two large epidemiological studies (15% by Guan et al, and 12.8% by Chinese CDC).^2,17^ This discordance could be explained by the age difference, the latter two studies had a median age of 47 and 48 years respectively, compared to 67 years from our report. COVID-19 patients with hypertension appeared to have a higher death rate and were more frequently seen in severe cases.^17^ We also examined the death rate and the incidence of critical cases in our cohort. Compared with non-hypertension controls, COVID-19 patients with hypertension had a higher death rate (10.3% [13 of 126]) *vs* 6.4 [eight of 125]) and incidence of critical cases (18.3% [23 of 126]) *vs* 11.2 [14 of 125]), but these differences failed to reach statistical significance. However, we did find that patients remained in hospital had a significantly longer disease course in COVID-19 patients with hypertension than those without (36.7±12.4 *vs* 31.6±7.7; p=0.024). Therefore, our results, together with those from other groups, provided ample evidence that hypertension is a critical risk factor for the poor clinical outcome of COVID-19 patients. Indeed, this notion was further supported by laboratory testing results. COVID-19 patients with hypertension had much lower blood oxygen index (p<0.001), as well as higher blood urea (p=0.020) and ALT (p=0.022) than those without hypertension.

The mechanisms by which hypertension results in the poor clinical outcome of COVID-19 remain obscure. We found that compared with non-hypertension controls, COVID-19 patients with hypertension have markedly increased levels of inflammatory hs-CRP (p=0.024), procalcitonin (p=0.017) and IL-6 (p=0.017), suggesting that dysregulated inflammatory response might contribute. In fact, hypertension has been well-known for its capacity in stimulating adaptive response and inducing the elevated production of inflammation cytokines.^25,26^ AT II, an effector peptide of the RAS, which is responsible for the pathophysiology of hypertension,^27^ has also been demonstrated to be capable of inducing the production of IL-6, IL-1β, TNFα, IFNγ, IL-17 and IL-23 in multiple animal models.^28^ Not surprisingly, treatment of hypertension patients with ACEIs and ARBs, which functional through reducing the level of AT II and increasing the expression of ACE2, effectively downregulated the production of inflammatory cytokines.^29^

Increased levels of inflammatory cytokines has also been observed in COVID-19 patients with or without hypertension as a coexisting illness. Huang et al reported that the plasma concentrations of IL-1β, IL-1RA, IFNγ, TNFα and other cytokines were significantly higher in both ICU patients and non-ICU patients with SARS-CoV-2 infection than in healthy adults. Further comparison found that IL-2, IL-7, IL-10, IP-10, MCP1, MIP1A, and TNFα were higher in ICU patients than non-ICU patients.^19^ Elevated concentration of IL-6 were also demonstrated in COVID-19 patients with severe and critical disease compared with those with mild and common illness.^30,31^ Based on these evidence, a multicenter, randomized controlled trial was recently registered on Chinese Clinical Trial Registry (ChiCTR2000029765) to evaluate the efficacy and safety of IL-6R blockade with tocilizumab in the treatment of COVID-19.

As mentioned above, inhibiting dysregulated inflammatory response represents a promising therapeutic strategy for both COVID-19 and hypertension patients. Therefore, ACEIs and ARBs that are capable of reducing the production of inflammatory cytokines are potential candidate drugs for treatment of COVID-19 patients with preexisting hypertension, as suggested by several groups.^12,32^ However, the clinical evidence is still missing. In this report, we found that in the 43 COVID-19 patients with hypertension treated with ARBs/ACEIs before and after diagnosed with SARS-CoV-2 infection, the concentrations of inflammatory CRP and procalcitonin were significantly lower compared with those treated with non-ARBs/ACEIs. ARBs/ACEIs treatment also resulted in a lower death rate and less critical cases in these patients. We did not find the concentrations of IL-6 in the ARBs/ACEIs group to be significantly different from that in the non-ARBs/ACEIs controls, which suggests that ARBs/ACEIs treatment alone might not be efficient in modulating the production of this cytokine. However, combining of ARBs/ACEIs and IL-6R blockade could presumably have a synergic effect in regulating the elevated inflammatory response in COVID-19 patients with preexisting hypertension, which could be tested in future clinical trials.

ARBs/ACEIs treatment has been reported to increase the expression of ACE2,^33,34^ which is also the cellular receptor for SARS-CoV-2 infection.^8^ Although the downregulation of ACE2 following SARS-CoV infection has been reported to result in acute lung injury, suggesting a protective role of ACE2 upregulation and ARBs/ACEIs treatment in COVID-19, it still raised concerns that ARBs/ACEIs treatment could facilitate SARS-CoV-2 infection by increasing the expression of ACE2.^12,35,36^ Therefore, we compared the frequency of ARBs/ACEIs usage in COVID-2019 patients with hypertension with those in hypertension patients admitted to HPHTCM before COVID-19 emerged, and found that they were not statistically different (34% [43 of 126] *vs* 35.4% [688 of 1942]; p=0.797). Our result thus indicated that ARBs/ACEIs treatment did not pose an added risk for SARS-CoV-2 infection in our study cohort. ACE2 was reported to bind to SARS-CoV-2 with approximately 10- to 20-fold higher affinity than that bind to SARS-CoV. This finding, together with the fact that people from different races, with different ages and sexes were all susceptible to SARS-CoV-2 infection, suggest that physiological expression of ACE2 may be already sufficient for SARS-CoV-2 infection, and further upregulation might not increase the risk.

ARBs and ACEIs are both antihyperhensive drugs discovered to maintain the blood pressure. Thus, ARBs/ACEIs treatment could result in hypotension in healthy subjects, which may prevent their application in COVID-19 patients without hypertension. Alternatively, novel therapeutic options using ACE2 as target will be promising in treating SARS-CoV-2 infection without affecting blood pressure.

Our study has several limitations. First, due to the retrospective nature of this study and the fact that it was conducted in a single hospital, interpretation of our findings might be limited by the sample size and selection bias. We also realize that there are additional risk factors may not be well-controlled despite as many confounders as possible were corrected for. Nevertheless, as far as we know, this is the largest retrospective cohort study designed to examine the usage of ARBs/ACEIs and its effect on COVID-19 patients with preexisting hypertension. Second, the severity and disease course were not identical among these patients, which resulted in the difficulty in collecting laboratory indicators at the same time point, we therefore selected the most extreme values beyond the normal range of laboratory indicators that could reflect the severity of condition for comparison. However, this strategy might still cause biases in presenting laboratory indicators. Third, the expression of ACE2 and other inflammatory factors were not determined in this study due to the sample availability and limited technical resources in our hospital, this limitation prevented us from further exploring the mechanisms by which ARBs/ACEIs regulate the inflammatory status of COVID-19 patients with hypertension.

## Perspectives

The evidence presented in this study supports the use of ARBs/ACEIs over other antihypertensive drugs in treating COVID-19 patients with preexisting hypertension. Large prospective studies are required to confirm this finding, and to explore the mechanisms by which ARBs/ACEIs regulate the inflammatory response and prevent disease progression. These efforts might eventually lead to the development of novel therapeutic agents targeting ACE2 to treat non-hypertensive COVID-19 patients without affecting blood pressure.

## Data Availability

After publication, the data will be made available to others on reasonable requests to the corresponding author. A proposal with detailed description of study objectives and statistical analysis plan will be needed for evaluation of the reasonability of requests. Additional materials might also be required during the process of evaluation. Deidentified participant data will be provided after approval from the corresponding author and Hubei Provincial Hospital of Traditional Chinese Medicine.

## Acknowledgement

We thank all patients and their families involved in the study, and all the healthcare professionals from Hubei Provincial Hospital of Traditional Chinese Medicine who helped and took care of the patients with COVID-19 for their great effort and selfless dedication in the medical relief operation against SARS-CoV-2.

## Sources of Funding

This work was supported by research grants from the National Natural Science Foundation of China (2017NSFC81670825 and 2020NSFC31970865). The funders had no role in study design, data collection and analysis, decision to publish, or preparation of the article.

## Conflicts of Interests/Disclosures

We declare no competing interests.

## Novelty and Significance

### What Is New?

- Hypertension has been reported to be the leading coexisting illness of COVID-19 patients, and these patients were frequently treated with ARBs/ACEIs to control their blood pressure.
- The effects of ARBs/ACEIs on the pathogenesis of COVID-19 has yet to be defined.

### What Is Relevant?

- We conducted the first and the largest case series to evaluate the effect of ARBs/ACEIs on the pathogenesis of COVID-19 patients with preexisting hypertension.
- We found that without increasing the risk for SARS-CoV-2 infection, ARBs/ACEIs outcompeted other antihypertensive drugs in containing exaggerate inflammatory response and curbing disease progression in COVID-19 patients with preexisting hypertension.

## Summary

Our results support the use of ARBs/ACEIs over other antihypertensive drugs in treating COVID-19 patients with preexisting hypertension. Large prospective studies are required to confirm this finding, and to explore the mechanisms by which ARBs/ACEIs regulate the inflammatory response and prevent disease progression.

## Notes

### Competing Interest Statement

The authors have declared no competing interest.

